# EDUCATIONAL NEEDS OF MEDICAL PRACTITIONERS ABOUT MEDICAL BILLING: A SCOPING REVIEW OF THE LITERATURE

**DOI:** 10.1101/2021.04.29.21256362

**Authors:** Margaret Faux, Jon Adams, Jon Wardle

## Abstract

**Introduction:** The World Health Organization has suggested the solution to health system waste caused by incorrect billing and fraud is policing and prosecution. However, a growing body of evidence suggests leakage may not always be fraudulent or corrupt, with researchers suggesting medical practitioners may sometimes struggle to understand increasingly complex legal requirements around health financing and billing transactions, which may be improved through education. To explore this phenomenon further, we undertook a scoping review of the literature to identify the medical billing education needs of medical practitioners and whether those needs are being met.

**Methods:** Eligible records included English language materials published between 1 January 2000 and 4 May 2020. Searches were conducted on MEDLINE, PubMed, Google Scholar, CINAHL, LexisNexis and Heinonline.

**Results:** We identified 74 records as directly relevant to the search criteria. Despite undertaking a comprehensive, English language search, with no country restrictions, studies meeting the inclusion criteria were limited to three countries (Australia, Canada, U.S), indicating a need for further work internationally. The literature suggests the education needs of medical practitioners in relation to medical billing compliance are not being met and medical practitioners desire more education on this topic. Evidence suggests education may be effective in improving medical billing compliance and reducing waste in health systems. There is broad agreement amongst medical education stakeholders in multiple jurisdictions that medical billing should be viewed as a core competency of medical education, though there is an apparent inertia to include this competency in medical education curricula. Penalties for non-compliant medical billing are serious and medical practitioners are at risk of random audits and investigations for breaches of sometimes incomprehensible, and highly interpretive regulations they may never have been taught.

**Conclusion:** Despite acknowledged significance of waste in health systems due to poor practitioner knowledge of billing practices, there has been very little research to date on education interventions to improve health system efficiency at a practitioner level.

## INTRODUCTION

The World Health Organization (WHO) has stated that “health-care systems haemorrhage money,” citing ten causes of inefficiencies and remedies.^1^ In the cited domain of waste attributable to fraud and corruption, the solutions proffered focus on measures to police and sanction wrong doers, such as medical practitioners who over-service in fee for service payment environments.^1^ Notably absent is any suggestion that teaching medical practitioners how their health systems work and how to allocate health dollars correctly may improve their compliance and reduce waste. This is despite evidence from the U.S, Canada and Australia suggesting medical practitioners may have at best, only a cursory understanding of the complex financial and billing infrastructure in their health systems, which may be contributing to unintentional misuse and exposure to serious legal sanctions.^2^

In Australia, despite an overarching assumption that doctors have high legal literacy in relation to correct billing using Australia’s national universal health system, Medicare,^2^ a recent study seeking to measure that experience, challenged that assumption, suggesting medical practitioners may instead be experiencing difficulties accessing reliable medical billing advice.^3^ In 2016, the Government of the Netherlands acknowledged this educational gap by introducing a requirement that universities and medical specialist training colleges provide education to medical practitioners in relation to medical billing and the costs of providing care, the stated aim being to tackle billing mistakes and fraud through prevention, rather than solely through punitive post-payment policing.^4^ While this intervention has been implemented, it does not appear to have been evaluated. However, the Dutch Healthcare Authority now details how consumers can report suspected healthcare fraud.^5^ This may suggest that successful implementation of medical billing education has placed the Netherlands Government in a better position to prosecute deliberate misconduct when it is reported.

However, while medical billing education has been recognised as an effective measure to improve compliance, reduce incorrect billing and improve integrity of health financing systems,^6^ formal education initiatives remain rare and many medical practitioners may have received no training whatsoever.^7^ To explore this phenomenon further, a scoping review of the literature was undertaken^8^ to determine the extent to which focused examination has been undertaken of the educational needs of medical practitioners in relation to medical billing compliance and whether those needs are being met.

## METHODS

### Search strategy and selection criteria

Inclusion criteria targeted literature that specifically cited teaching and education of medical practitioners in relation to medical billing, using combinations of keywords such as “medical billing and education”, “medical billing and curricul*”, “billing and coding education”, “physician medical billing”, “Medicare billing education”. The word “coding” was included in the keywords, because medical billing is referred to as medical coding in some jurisdictions. Materials dealing with individual health care system specifics and medical billing in the broad contexts of health economics, politics and health policy were deemed not relevant and excluded.

Grey, commentary materials and legal literature were included in the search strategy and manual searching was undertaken to review bibliographies and reference lists in the material originally sourced. No country restrictions were put in place.

As this is a novel topic and of interest to the general health, social sciences and legal communities, relevant databases in these areas were initially searched including MEDLINE, PubMed, Google Scholar, CINAHL, LexisNexis and Heinonline. We initially included the CINAHL nursing and allied health database, to capture possible results from multi-disciplinary billing settings such as Rehabilitation Medicine and Palliative Care. However, no relevant results were returned so CINAHL was later excluded. LexisNexis and Heinonline are important legal databases, which were included as they are likely repositories of law reports and articles dealing with medical practitioners who had been prosecuted for incorrect billing through law enforcement, as the WHO recommends. In countries where the rule of law is upheld, education about laws is usually made available prior to individuals being required to engage with those laws. We therefore searched these databases to determine whether medical practitionres had discussed educational needs in the context of policing and prosecution for incorrect billing. LexisNexis returned numerous irrelevant results which were unable to be reduced by refining search terms. All results found on LexisNexis were duplicates of those found on Heinonline and due to Heinonline enabling more granular refinement of search criteria, we excluded the LexisNexis database in final searches.

Due to the large number of initial search hits, numerous filtering strategies were applied and criteria refined until sensitivity and specificity appeared to be optimised. This process identified 3022 records of materials published in the last 20 years. We undertook further manual searching on Google Scholar to ensure any grey literature were found as well as again manually reviewing bibliographies and reference lists in the material originally sourced.

As this topic tends to divide opinion along partisan lines (i.e. “medical practitioners are deliberately committing fraud”, or, “are unintentionally making errors”), opinion pieces and grey literature had the potential to be very relevant in the evolving discussion on the causes of non compliant medical billing. To ensure we did not reject key insights numerous government reports were included. Only two empirical Australian studies directly related to the research topic were found.

## RESULTS

After removing duplicates and unrelated records, we screened the abstracts of the remaining 241 records, and excluded a further 155 records which did not meet inclusion criteria, because they did not specifically target educational needs of medical practitioners around medical billing. We also excluded a further 12 records which were legal cases concerning non-compliant medical billing and fraud, because they did not specifically address teaching and education of the medical practitioners who were the subject of those proceedings. An additional 44 records met the inclusion criteria as a result of manual processes. The majority of relevant results on medical billing in Australia were found in grey literature and commentary, which may therefore have an inherent bias. While in the U.S the topic appears to be more mature, with substantial numbers of empirical studies found. In Canada, only one empirical study and one commentary article met the inclusion criteria. Summary results of the search are presented in Table 1. Although a comprehensive international, English language search with no country restrictions was conducted, results were limited to three countries (Australia, Canada, U.S). The final results were sorted into four categories, presented in Table 2.

**Table 1.**
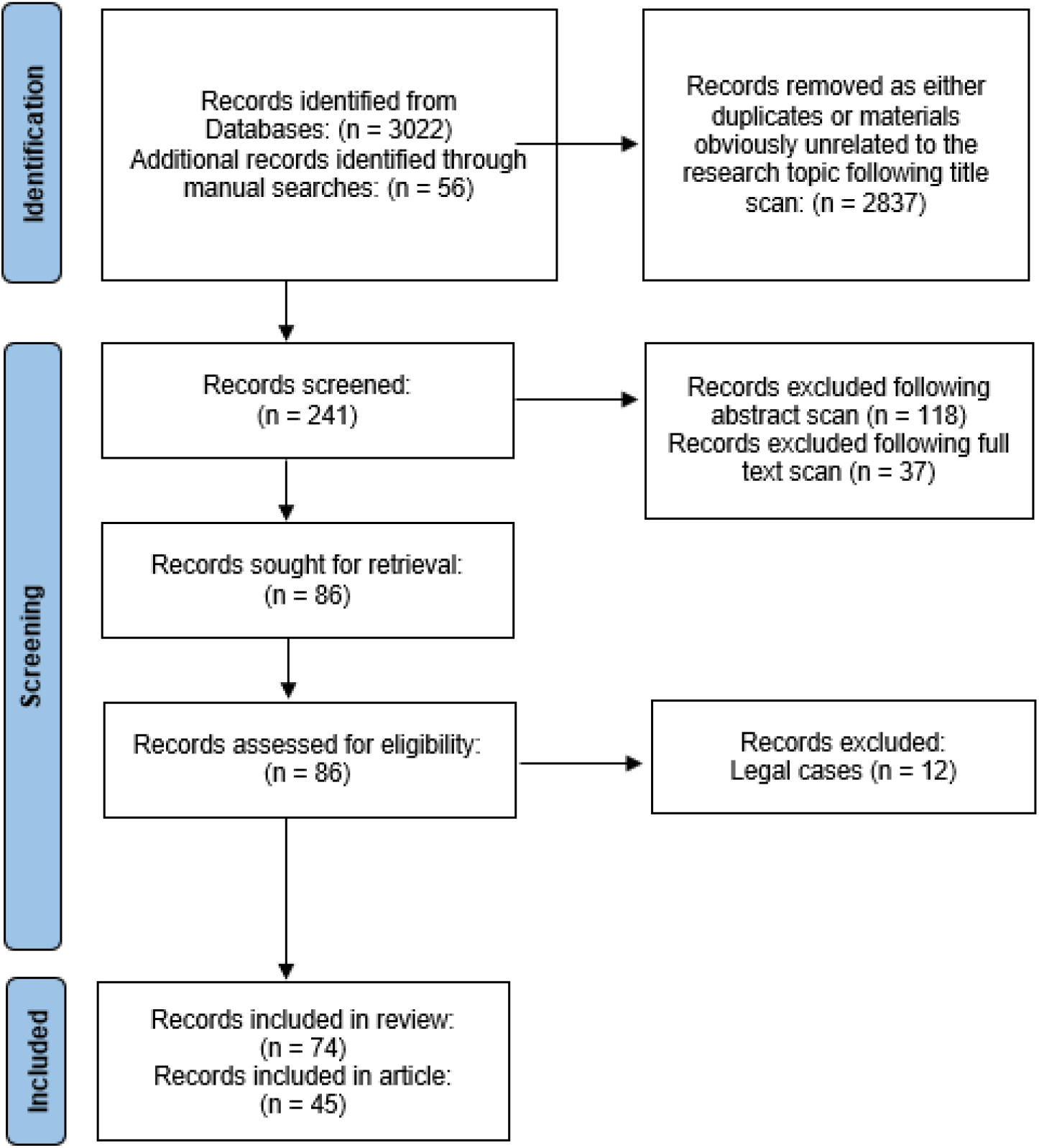
Prisma Flow Diagram.

**Table 2.** Final Search Results.

| Category | Australia | U.S | Canada | Total |
| --- | --- | --- | --- | --- |
| Empirical | 2 | 28 | 1 | 31 |
| Grey | 37 |  |  | 37 |
| Commentary/opinion | 3 | 2 | 1 | 6 |
| **TOTAL** |  |  |  | **74** |

For ease of reference, what follows is a stepwise presentation of the results in Table 2, commencing with empirical literature and ending with commentary and opinion pieces.

### Empirical literature - Australia

A 2004 doctoral thesis on the topic of Medicare fraud and inappropriate practice provided a detailed analysis of how fraud and overserving allegedly became entrenched in Australia’s health system between 1975 and 1996.^9^ The study was “primarily an empirical study” which included over 59 qualitative interviews with politicians, leading stakeholder representatives, senior public servants, fraud investigators, journalists, and others who spoke on condition of anonymity. The study suggested the extent of non-compliant medical billing in Australia at the time may have been over 25% of the schemes’ total cost, and definitely not under 10%. Precise quantification was not possible. Solutions to non-compliance were positioned through a criminal justice lens, with education only briefly mentioned as a weaker, less effective solution than regulation and policing. The thesis argued lax regulation and inadequate resourcing had led to a failure of necessary oversight and prosecution of errant medical practitioners.The study did not offer any explanation for non-compliant medical billing beyond deliberate abuse, and most interview participants appeared to share the view that medical practitioners “know how to bill correctly…” though subsequent research suggests this may not be the case.^3^

In a study of medical practitioner education stakeholders^3^ the authors conducted a national cross-sectional survey which reported the first attempt to systematically map the ways Australian medical practitioners obtain education and understanding of medical billing, and explored the perceptions of medical education stakeholders on the topic. The results revealed little medical billing education was occurring with the majority of participants (70%, n=40) reporting they did not offer and had never offered medical billing education. However, 89% of participants thought medical billing education should be provided but there was no consensus on who should provide it or when it should occur. The study also found that most education in this area occurs on an ad hoc basis and is taught by medical practitioners who themselves have never been formally taught correct use of the Medicare scheme because no national, government approved curriculum has ever existed. The knowledge of those teaching the topic was therefore reported as variable, and the researchers reported this as being **c**onsistent with U.S findings, which suggest that rather than reliance on ad-hoc training, development of a national medical billing curriculum should be encouraged to improve compliance, expedite judicial processes and reduce waste.

### Empirical literature - U.S and Canada

Our review found studies specifically seeking to measure an equivalent experience have been primarily undertaken in the U.S, where a different medical billing system to Australia’s operates, and where the heterogeneity of service providers and payers may warrant additional focus on billing education. The Australian medical billing system is based on a unique schedule of service codes known as the Medicare Benefits Schedule (MBS), whereas the U.S uses the International Classification of Disease (ICD) and Current Procedural Terminology (CPT) codes. Canada uses different billing codes again, known as the Ontario Health Insurance Plan (OHIP) Schedule of Benefits and Fees. However, an assessment of the differences between these code sets and the practical application of each suggests the challenges faced while undertaking medical billing in all three countries is similar because the cognitive process of matching clinical encounters to an administrative dataset is the same.

U.S research on the subject of medical billing and reimbursement is more advanced than in Australia due to increased recognition in that jurisdiction that medical billing is a component of every interaction between a patient and a medical practitioner.^7, 10–16^ The U.S literature suggests that training in the area of medical billing should be viewed as a core competency and a national curriculum on the topic should be developed.^7^ However, despite the Accreditation Council for Graduate Medical Education in the U.S agreeing that education about practice management and economics forms part of the required core competencies for medical practitioners, teaching of those subjects is variable and no formal national curriculum exists. One of the recognised challenges identified in the U.S material is that of ‘teaching the teachers.’^7^ With no written curriculum on the topic of medical billing, researchers pointed out that teaching of the subject will be variable and will depend on the expertise, experience and the confidence of senior mentors who may themselves have had little training in the area.

In one study involving a cross sectional, needs assessment survey of second year community and university based internal medicine residents from four U.S geographic regions,^10^ participants (n=133) completed a questionnaire which included 27 questions, and the findings indicated that medical practitioners rated their own knowledge of Medicare billing as low. Participants also strongly agreed that their training in medical billing was inadequate and that it was important and should be a requirement of residency training programs.

In a 2009 study examining the adequacy of training in the area of medical billing and coding as perceived by 2300 recently graduated paediatricians recruited from the American Board of Pediatrics database of recent graduates^7^ less than 20% of respondents reported their training in medical billing and coding as adequate. The key points emanating were that medical billing and coding is not uniformly taught and should be included in the core competency requirements for medical residents. Further, that work needs to be done to develop and test a curriculum in medical billing and coding and that residency programs need to ensure they are equipped to practice.

In another descriptive study of 104 medical students examining attitudes to professionalism,^11^ preferences in the importance of professional competencies, teaching preferences in professionalism and the egregiousness of 30 vignettes of professional misconduct, participants rated illegal billing as the second most egregious of 30 vignettes of misconduct. Substance abuse was reported as being the most serious misconduct (86.8%), followed by illegal billing (69.1%) which was rated higher than sexual misconduct (50%).

Since 2016, we found an increase in the number of U.S studies on this topic, where results have echoed earlier findings that the level of medical billing literacy amongst medical practitioners remains demonstrably low and may be improved by targeted education.^11–15^ In one recent U.S study more than 70% of medical practitioner participants felt there was a need for medical billing and coding to be included in the medical curriculum^16^ and a 2019 study of senior residents and staff physicians in Ontario, Canada (n=33)^17^ described the billing accuracy of the medical practitioner participants as poor overall, with billing errors and omissions causing substantial revenue losses. Participants in that study felt that current medical billing education was both insufficient and ineffective and desired more.

### Grey Literature and Commentary

A review of policy and parliamentary papers uncovered numerous Australian government reports dealing with medical billing compliance, and a 2018 analysis and critique of the U.S government’s approach to managing Medicare compliance mirrored many of the challenges being experienced in relation to medical billing compliance in contemporary Australia.

### Government reports – Australia

The principle government reports uncovered were the Annual Reports of the Professional Services Review Scheme (PSR) in Australia.^18^ The PSR was established in 1994 as a peer review scheme to investigate Medicare services billed by medical practitioners, with the objective of protecting the integrity of the scheme.

A review of 25 years of the annual reports reveals the PSR has been plagued by costly legal challenges by medical practitioners who have felt aggrieved by a lack of due process, flawed extrapolation methodologies and inadequate legal reasoning to support adverse findings against them. The annual reports also consistently cited medical practitioner confusion about correct billing practices. Unfortunately, full decisions of the PSR, which may assist practitioners to understand how to bill correctly have never been published due to codified secrecy provisions which protect the agency from public scrutiny.

The operation of the PSR was the subject of a Senate Enquiry in 2011.^19^ During the enquiry, submissions from medical practitioners highlighted both the complexity of Medicare billing and the inadequacies in the resources available to them concerning its proper use. This directly contradicted institutional submissions from Medicare suggesting that ample resources and reliable support were available. One submission by a medical defence union representative indicated that processes should be in place to enable medical practitioners to obtain clarity about the use of the MBS and another drew a comparison between the advice and written rulings available from the Australian Taxation Office and the lack of such information and advice from Medicare, suggesting that this meant medical practitioners could unknowingly fall into error. The Senate Committee concluded that, although it was the responsibility of medical practitioners to make clinical judgments, as much advice and information as possible should be available to them in relation to MBS itemisation. However, the committee was silent as to who should provide this advice and information.

In addition to the PSR reports and Senate Enquiry, manual searches revealed a departmental newsletter to the profession in February 2007 titled ‘Education the Key to Compliance’ in which the government announced that by changing medical practitioner claiming and prescribing behaviour through an education and compliance program, $250 million in Medicare program savings had been achieved in the previous year.^6^

### Commentary on the Medicare appeals process - U.S

The challenges plaguing the Australian PSR appear similar to those reported in the U.S, where one commentator described the U.S Medicare appeals system as broken,^20^ and a U.S court has pondered whether Medicare laws have become so byzantine that the government had lost control of them.^20^ The combined effects of complex, constantly changing, opaque medical billing rules and the use of extrapolation techniques appear to be at the heart of the problem which may have rendered the U.S government unable to manage medical billing compliance under its fee-for-service Medicare scheme, to the point where it “seems unable to keep up with it’s own frenetic lawmaking.”^20^ Further, that the U.S Department of Health and Human Services conceded it would take more than 10 years to clear the backlog of Medicare appeals awaiting review by an Administrative Law Judge (ALJ) noting ALJs overturn decisions against medical practitioners over half of the time.^20^ This may suggest that like their Australian counterparts, U.S medical practitioners may be struggling to understand complex medical billing rules they have never been taught and appearing before an ALJ is the first time they are afforded a merit based, evidentiary hearing and benefit from due process before a truly independent arbiter.

### Canadian commentary

A recent publication in the *Journal of Medicine and Philosophy* initiated an important discussion concerning the moral dimensions around compliant medical billing, suggesting creative billing practices should be stigmatized rather than celebrated from within the profession itself.^21^ The author described as a ‘rather surprising oversight’ that while medical ethics is a recognized component of medical education, the financial aspects of medical practice are almost never discussed and medical practitioners therefore receive little or no guidance in this important area. Further, that in fee-for-service payment environments, medical practitioners have enormous latitude in regards how they describe their services, with often very little effective oversight by payers. Therefore, the human temptation to misrepresent the services they provide can sometimes be significant, particularly where a seemingly small ‘fiddle’ to a service description can lead to higher reimbursement. The related ethical challenges are never taught nor mentioned throughout medical undergraduate or postgraduate training, yet the legal consequences when medical practitioners are found in breach of payment rules are usually very serious. The author argued that both medical schools and specialist colleges have failed in their duty to address this critical gap in learning and suggested some colleges may actually be cultivating the practice of questionable or borderline billing to ‘maximise’ or ‘optimize’ financial return. Moreover, that medical practitioners often fail to see the connection between their own poor billing conduct and the failure of the health system overall and that to address these challenges, both education and regulation are required.

### Australian government educational materials

We found a number of resources produced by Medicare described as ‘Compliance Education for Health Professionals.’^22^ These include a “Medicare Billing Assurance Toolkit” and various e-learning modules. A review of these resources found a heavy focus on penalties for non-compliance without providing comprehensive information on how to be compliant. The resources suggested an overarching departmental view that medical practitioners possess a high level of legal literacy regarding correct use of Medicare, though available evidence challenges this position.^3^ The resources were found to be rudimentary, offering little more than directing medical practitioners to the MBS if they are unsure of billing requirements, which is unhelpful considering findings of a recent study suggested the MBS has become complex and incomprehensible.^2^

Where education does exist, it may not be directed to the relevant parties. During manual searching from the bibliographies and references lists in the preliminary searches, a training course was found that appears to be the only government accredited course in Australia dealing with the processing of medical accounts.^23^ On review of the course materials, performance criteria and outcome measures, it was found that this was a basic certificate level course designed for medical receptionists who are not responsible for MBS billing, rather than being targeted at medical practitioners who are.

### U.S government educational materials

We also reviewed educational materials available to U.S medical practitioners who we found are similarly required to self-learn the complexities of medical billing by reading a number of resources such as Explanation of Benefits Remittance Statements they receive when the claims they submit are denied, publications produced by intermediaries in the medical billing process who are contracted by the federal government (known as Medicare Administrative Contractors), and materials on the Centres for Medicare and Medicaid Services website.^23^ However, evidence suggests medical billing literacy among U.S medical practitioners remains low^7, 10–16^ and the above resources are inadequate to prepare them to bill correctly and protect them from post-payment investigation.

## DISCUSSION

The legal machinery underpinning fiscal transactions in health systems is typically profoundly complex. We found that the paucity of available data on this important topic does not correlate well with the impact non-compliant billing has on global health systems. Irrespective of whether the cause of non-compliance is deliberate or accidental, the size of the problem, which has been reported as averaging 7% of total health expenditure,^25^ is of sufficient magnitude to warrant focussed academic attention, particularly given the likely global economic slowdown caused by Covid-19. Waste caused by non-compliant medical billing in health systems can no longer be ignored. The fact that the scope and extent of this issue as a problem has been consistently identified as a major barrier to the efficiency of health systems, yet few studies have been conducted on initiatives that may help to address the issue, suggests that further research is warranted to ensure that stakeholders are able to make evidence-informed decisions when developing initiatives to combat medical billing non-compliance.

Although limited to three countries, the literature revealed a pervasive unified global view across those countries that medical practitioners obtain high levels of medical billing literacy through an osmotic process unsupported by the evidence. Unmet education needs were also evident throughout the literature across jurisdictions. Early reports^9^ uncoveed by our review mention short term success with education initiatives for medical practitioners in Australia, and the PSR consistently cited practitioner confusion as being an ongoing problem. However, from the outset, very little was published in the PSR annual reports concerning available assistance to medical practitioners concerning how to use the (Australian) Medicare system correctly. The first PSR Director repeatedly advised medical practitioners via these reports to ‘read the MBS book each year’ and suggested speaking with Medicare when unsure of correct itemisation. However, this is and was always an unrealistic and onerous requirement on medical practitioners given the current printed version of the MBS comprises over 900 A4 pages of item numbers, explanatory notes, rules and cross references, many of which are difficult to comprehend, and a single medical service can be the subject of over 30 different payment rates and rules.^26^ We also found no evidence of medical billing educational resources such as a 25 year body of precedent that might assist medical practitioners to understand how to bill correctly.

In the U.S, government maladministration was described as having far reaching consequences impacting the broader health system and ultimately consumers,^20^ and we suggest the impacts identified would be applicable in any health system.

The first such impact is that medical practitioners, as small business owners, may not have the financial means to support lengthy investigations and repay large amounts, so may become insolvent or choose to stop practicing. This causes the health market to contract to the detriment of smaller providers and their patients, becoming consolidated by larger corporations with the liquidity to withstand long legal battles. Further, if small providers servicing remote communities are impacted, their absence may not be filled by larger corporates, potentially leaving such communities without medical services.

A second impact was cited as regulatory and administrative burdens causing some medical practitioners to stop treating Medicare patients completely. In Australia, where all citizens and many eligible residents are covered by Medicare, the practical expression of this type of pressure is seen when medical practitioners simply stop engaging with Medicare directly, requiring patients to instead pay full fees upfront and claim available rebates themselves. This practice is evident in the current out-of-pocket medical fees crisis in Australia.^26^

Another serious and potentially dangerous impact is that working under the constant threat of audits and investigation may cause some medical practitioners to under-service their patients. Others may continue to provide services but not bill and be reimbursed for them, reducing government visibility over actual service delivery. A recent study in Australia described evidence of such practices among medical practitioners.^28^

Medical practitioners act as stewards for the integrity of their health systems through the bills they submit for each clinical encounter.^2^ Further, medical billing is a component of every clinical encounter (whether directly or indirectly) and the penalties for non-compliance across jurisdictions are severe. Yet medical practitioners appear to receive little formal preparation in the proper use of the billing and payments systems they are required to engage with. Moreover, opaque and interpretive medical billing codes cause difficulties for medical practitioners in multiple jurisdictions, yet no research has ever sought to examine how, when and where medical practitioners obtain the high levels of medical billing literacy expected of them.

Successful health financing systems depend on the fast flow of payments between patients, payers and providers in a context of high volumes of small transactions, often sourced from public money. For this reason, a high level of scrutiny is required to ensure the integrity and sustainability of such schemes. However, in achieving this, a proportionately high level of precision must be maintained in the area of service descriptions and billing rules, to protect the providers who often have no option but to engage and claim reimbursements.

## LIMITATIONS

A limitation of this review is the fact that results were drawn only from three countries, which may limit the generalisability of results. However, we view this as in important finding in-and-of itself, suggesting an urgent need for further work on this topic in other settings. The relatively large body of work from Australia may be reflective of the significant government role in Medicare, which unlike many universal health care systems is still reliant on fee-for-service provison by private providers, resulting in increased public accountability and interest in the topic in that country. Further work is required to examine the topic in other countries. Extensive investigation of informal, ad hoc, and spontaneous educational initiatives that may exist in some jurisdictions were not captured by this review and may be deserving of focussed research attention

## CONCLUSION

Despite the increased research outputs on this topic in recent years there appears to remain a mistaken global view, unsupported by scientific evidence, that medical practitioners naturally know how their health systems work and how to bill correctly, and that punitive measures are therefore the sole solution to waste caused by non-compliant billing practices. This is despite a growing body of evidence suggesting education may be effective in addressing this problem.

Emerging health systems can learn from the experiences of the health systems reported in this study by prioritising curriculum development in health financing law and practice. Educating medical practitioners about the operation of their health financing systems and how to allocate scarce health dollars correctly protects them from exposure to potentially serious legal consequences for non-compliance, and may improve the efficient and equitable distribution of national health budgets.

## Data Availability

The datasets used and/or analysed during the current study are available from the corresponding author on reasonable request

